# Premature Ventricular Contraction-Mediated Ventricular Fibrillation: Clinical characteristics, Application of Machine-Learning Algorithm and Outcomes of Catheter Ablation: Multicentric Case Series

**DOI:** 10.1101/2025.10.06.25337458

**Authors:** Abhishek Maan, Eric Stanton, Matthew Greydanus, Avdhesh Mann Btech, Ryan Van De Leur, Moneeb Khalaph, Philip Sommer, Neal Chatterjee, Mustapha El Hamriti

## Abstract

**Background:** Premature ventricular contractions (PVCs) are common in patients with and without structural heart disease. In a subset of patients, PVCs are associated with malignant ventricular arrhythmias including ventricular fibrillation (VF). There are limited clinical tools available to identify which patients with PVCs are at risk for VF.

**Methods:** We analyzed data from an international, multi-center cohort of 61 patients who underwent catheter ablation for PVCs (41 with PVC-triggered VF, 20 controls). We evaluated the prevalence of routine 12 lead ECG characteristics in patients with PVC-triggered VF including (a) early repolarization (ER) in inferior/ lateral leads and (b) QRS notching of the sinus beat or PVC. We evaluated whether a machine learning (ML) ECG algorithm (Factor ECG) could discriminate between individuals with PVC-triggered VF and individuals with PVC and no history of VF. Explainability analyses were performed to identify which components of the ECG waveform were associated with risk prediction.

**Results:** In 41 patients with PVC-triggered VF, there were a median of 8 ICD shocks/per patient prior to index PVC ablation. The mean coupling interval of the PVC to the antecedent sinus beat was 313±130 ms. When compared to controls, early repolarization (39% vs. 20%) and QRS notching (71% vs. 25%) were significantly more prevalent in individuals with PVC-triggered VF. After a median ablation of 1 [IQR: 1-3]), 82% of patients remained free of VT/VF and ICD shocks over a median follow up of 400 [90–2490] days. The ML ECG algorithm demonstrated reasonable discrimination of patients with PVC-triggered VF compared to PVC without VF (AUROC 0.85 [0.56-1.0]). Anterior ST segment deviation and left bundle branch like delay of the ECG waveform were salient contributors to ML-based prediction.

**Conclusions:** In patients with PVC-triggered VF, routine ECG parameters including early repolarization and QRS notching were present in up to two-thirds of patients and were more prevalent compared to individuals with PVC without VF. An ML-based ECG algorithm effectively distinguished between PVC-triggered VF compared to PVC without a history of VF.

## Introduction

Although the occurrence of premature ventricular contractions (PVCs) is relatively common [1] and often considered benign in otherwise healthy individuals without structural heart disease [2], there is an emerging recognition of malignant risk in a subset of patients who develop PVC-induced ventricular fibrillation. For example, in the acute and subacute setting after myocardial infarction, PVCs can trigger ventricular fibrillation (VF) and catheter ablation of these triggering PVC can be an effective tool to mitigate risk of ventricular arrhythmias [3, 4]. PVC triggered VF has now been recognized in the absence of structural heart disease and underlies the diagnosis of idiopathic VF [5]. Short-coupled PVCs and those originating from the His-Purkinje system have been identified as triggers for VF in patients with previous myocardial infarction [6] and their role has also been described in patients with inferolateral -J wave syndrome [7, 8]. These findings underscore the hypothesis that the mechanism of VF, more broadly, may be an interaction between vulnerable substrate and triggering PVC. Previous work in patients with idiopathic VF has generally centered on the triggering PVC including modes of initiation and coupling intervals between PVC and sinus rhythm [9]. Presently lacking, however, are clinically actionable strategies to identify vulnerable substrate and risk of PVC-induced VF in individuals with PVCs.

Previous studies in the general population have suggested that some 12 lead ECG characteristics such as early repolarization and QRS notching may be generally associated with a higher risk of SCD [10, 11]. Recent work has demonstrated that deep learning analysis of unstructured ECG waveforms may identify salient signatures of incident cardiovascular risk including, for example, the risk of PVC induced cardiomyopathy [12]. Whether such a deep learning approach could identify salient substrate in those with PVC-triggered VF is unknown but could provide a novel diagnostic tool to inform risk and management strategies. Therefore, in this study, we evaluate the incidence of ECG abnormalities (early repolarization pattern, QRS notching) in patients with PVC-triggered VF. We examine the predictive accuracy of a deep learning ECG algorithm (FactorECG) for PVC-triggered VF compared to PVC without a history of VF. Finally, we perform explainability analyses of the deep learning algorithm to identify which components of the ECG are most related to prediction of VF in individuals with PVCs.

## Methods

### Study population

The retrospective cohort included patients from an international (USA, Germany), multi-center registry of individuals undergoing catheter ablation for PVC-triggered VF between November 2018 and September 2024. A total of 61 patients were included in the analytical cohort (41 individuals with PVC-triggered VF and 20 controls). We excluded patients with a diagnosis of early repolarization syndrome (ERS), Brugada syndrome (BrS) and those with polymorphic ventricular tachycardia degenerating into VF. In addition, we excluded patients in whom VF was noted in the setting of active myocardial ischemia. The control group was comprised of patients (N=20) undergoing catheter ablation for symptomatic PVC and without a history of sustained ventricular arrhythmias during the study period.

### EKG analysis

We analyzed data from the baseline pre-ablation EKG (either in sinus rhythm or with clinical PVCs). To assess the presence of repolarization abnormalities and notching of the QRS complexes in sinus rhythm or that of PVCs, the EKGs were independently assessed by both AM and ES and both reviewers were blinded to clinical characteristics including classification of PVC with or without VF. Discordance in assessment was resolved by an independent third reviewer (MG).

Based on previously published literature, notching of QRS complex may represent heterogeneity in electrical conduction and regional conduction delay [13, 14] and could also be a sensitive marker for myocardial scar [15]. It has also been postulated that subtle disruptions in depolarization are best detected in lateral leads and more prominent disruptions are best seen in inferior leads. Therefore, we focused on assessment of QRS notching and early repolarization (ER) patterns in these leads. On the 12-lead EKG recorded at 25 mm/sec, we assessed the following abnormalities in both patients :

(a): Early repolarization (ER) pattern in at least 2 contiguous leads either inferior or the lateral leads. ER was characterized as J-point elevation in keeping with the guidelines of consensus statement [16]
(b): Notching or fragmentation of QRS complex of either the native sinus beat or the PVC complex [11] For the assessment of notching or QRS fragmentation, we adjudicated it to be positive finding only if it was present in 2 contiguous leads.
(c): Coupling interval of PVCs: this was defined as the time-interval between the end of QRS complex of the preceding sinus beat and the onset of the PVC.
(d): Duration of QRS interval of PVCs: as the interval measured from beginning to the end of QRS interval

### Machine-Learning algorithm based on explainable deep-neural network

Deep learning ECG analysis was performed using Factor ECG, a variational auto-encoder, the derivation of which has been previously published [17]. The original FactorECG model was trained on 1.1 million EKGs and had used a variational auto-encoder consisting of 3-parts: an encoder, the FactorECG which consisted of 32 continuous factors and a decoder which reconstructed EKGs using these 32 factors. After training, the encoder can be used to convert any median beat EKG into the FactorECG format [17, 18]. The FactorECG algorithm can be accessed using the hyperlink: (https://pln.ecgx.ai).

### Pre-Ablation imaging

All patients who underwent PVC ablation had either an echocardiogram or a cardiac MRI at baseline. The decision and selection of imaging modality prior to PVC ablation was driven by operator’s discretion.

### Ablation procedure

We also collected electroanatomic mapping data from all the patients who underwent ablation for the clinical PVC which was concluded to be the trigger for PVC-VF. During ablation, the PVC triggering VF was targeted for ablation as the clinically relevant PVC. For induction of PVCs, a combination of burst pacing and isoproterenol infusion was used. Activation mapping was performed if PVCs were present. If a triggering PVC could not be induced, then pace-mapping was used to aid in catheter ablation. If elimination of the clinical PVC triggering VF was not possible, then we adopted a substrate modification approach for denetworking of Purkinje network as the primary ablation strategy [19].

### Follow-up

Follow-up data on clinical outcomes and peri-procedural complications were collected. Specific data on recurrence of ventricular arrhythmias were collected from a combination of EKGs, interrogations and remote transmissions from ICDs on the follow-up clinic visits.

### Statistical analysis

We compared continuous and categorical variables between the two groups of patients using the student’s *t* test and Fisher exact test respectively. The prevalence of EKG abnormalities (ER pattern and notching of QRS) and location of PVC ablation sites during ablation is described in percentages. For the assessment of 32 factors of FactorECG and which factors encoded the most relevant information, we logistic regression models. To assess the performance of FactorECG as a diagnostic model to discriminate between PVC-triggered VF and PVC without VF, we used the receiver-operator characteristic (ROC) curve to calculate c-statistic. All the analyses were performed using STATA software (STATA Corp, College Station, Texas).

## Results

In our multicenter study, a total of 41 patients underwent catheter ablation for PVC triggered VF. The median number of ICD shocks received prior to catheter ablation was 8 [IQR: 2-60] per patient. Approximately one-third of patients (N=14, [34%]) presented with electrical storm prior to undergoing catheter ablation. On the pre-ablation 12-lead EKG, the majority of patients with PVC-triggered VF (30 of 41 patients (73%) had a right bundle branch block PVC morphology. When compared to control patients, the coupling interval of the PVC to the antecedent sinus beat was significantly shorter in the PVC-triggered VF group (313 ± 130 vs. 455 ± 48.07 msec, p < 0.001). The summary of comparison of baseline characteristics between the two group of patients is presented in **Table 1**.

**Table 1:**
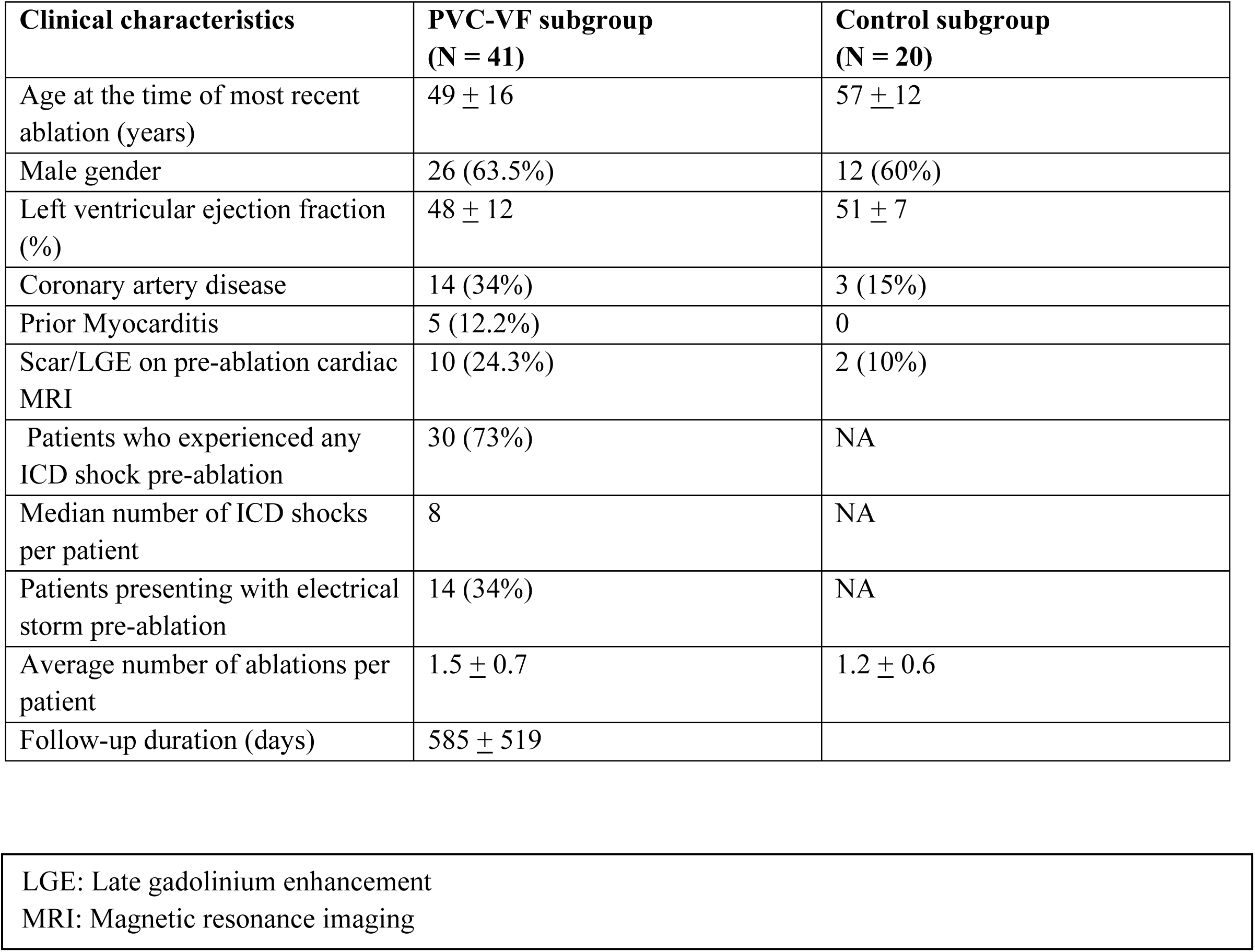
Comparison of baseline characteristics of patients who had PVC triggered Ventricular Fibrillation.

We next evaluated the prevalence of 12 lead EKG abnormalities in those with PVC-triggered VF compared to individuals with PVC without VF. We found that 39% of individuals with PVC-triggered VF had ER pattern in the inferior of lateral leads compared to 20% of individuals with PVC without VF (p = 0.001). Similarly, individuals with PVC-triggered VF had a higher prevalence of QRS notching as compared to control patients (71% vs. 25%, p = 0.001) (**Figure 1**).

**Figure 1:**
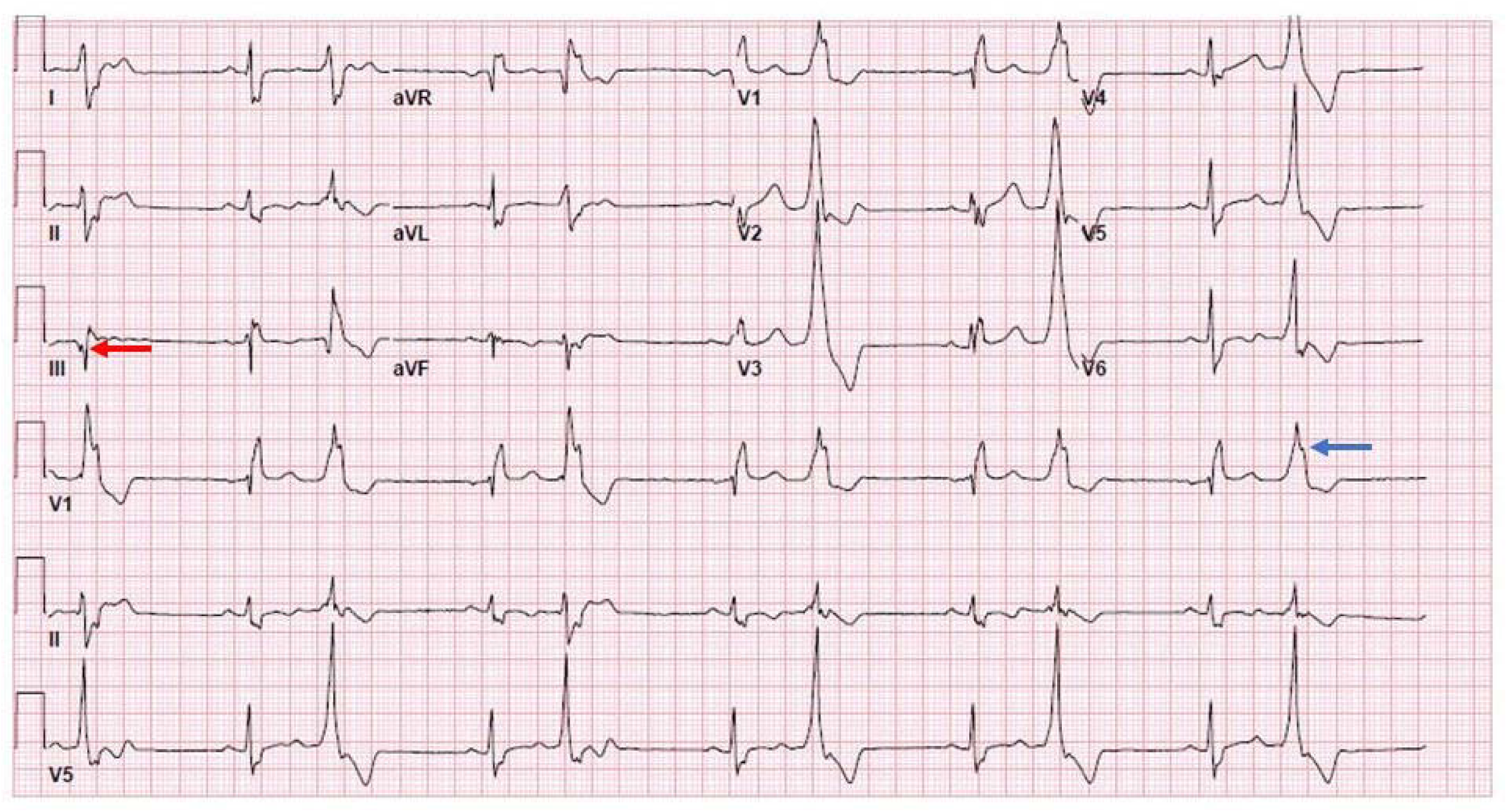
Figure showing notching of Sinus rhythm QRS complex (red-arrow) and notching of PVC complex (blue-arrow) in a patient presenting with PVC triggered Ventricular Fibrillation

### Ablation findings and clinical outcomes

During catheter ablation in patients with PVC-triggered VF, the median number of PVCs that were targeted was 1 [IQR: 0-4]. An ablation approach aimed at either targeting or de-networking of the Purkinje potentials was performed in two-thirds of these patients (61%) (**Figure 2 and 3**) The predominant location of PVCs in the PVC-VF group of patients was in the left ventricular septum (61%), followed by the left fascicular system and papillary muscles (12% each) (**Figure 4**). Non-inducibility of PVC-triggered VF as an acute procedural endpoint was achieved in all patients. The results of EKG features and ablation findings are summarized in **Table 2**. After a median of 1 [IQR: 1-3] ablation procedure, 83% of the patients (34 of 41) remained free of VF over a median follow up period of 388 [IQR of 42-2490) days.

**Figure 2:**
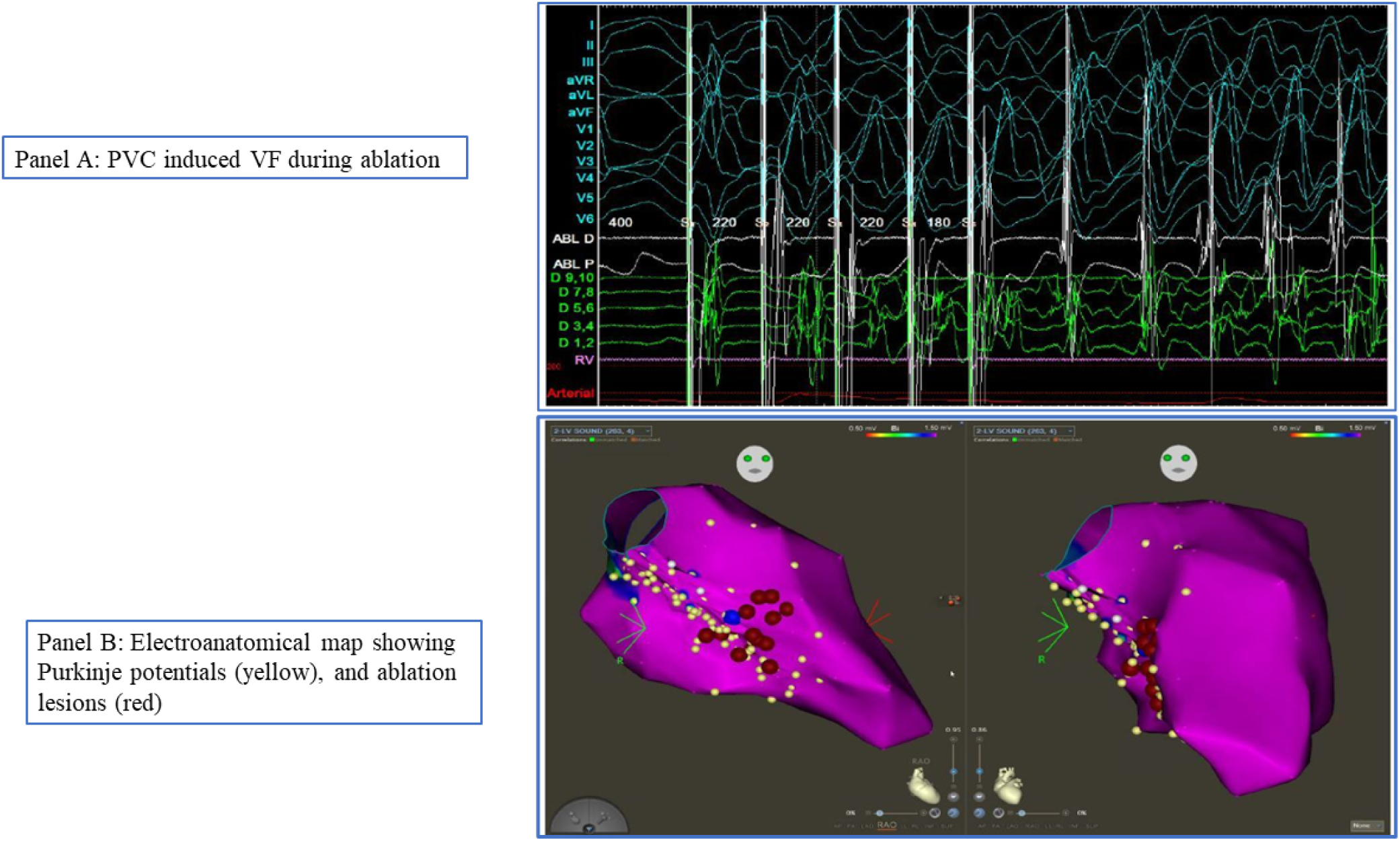
Figure showing PVC induced VF and ablation targets (Purkinje potentials) during PVC triggered VF ablation.

**Figure 3:**
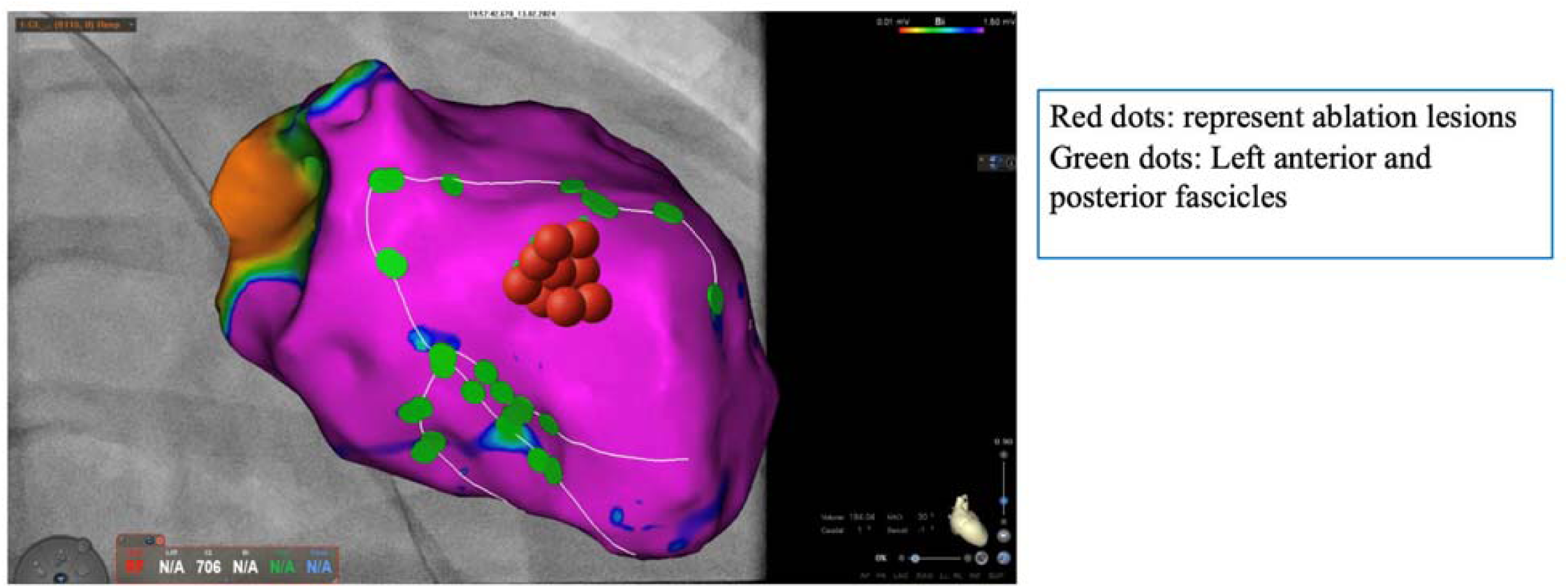
Figure showing ablation lesions and left anterior and posterior fascicles.

**Figure 4:**
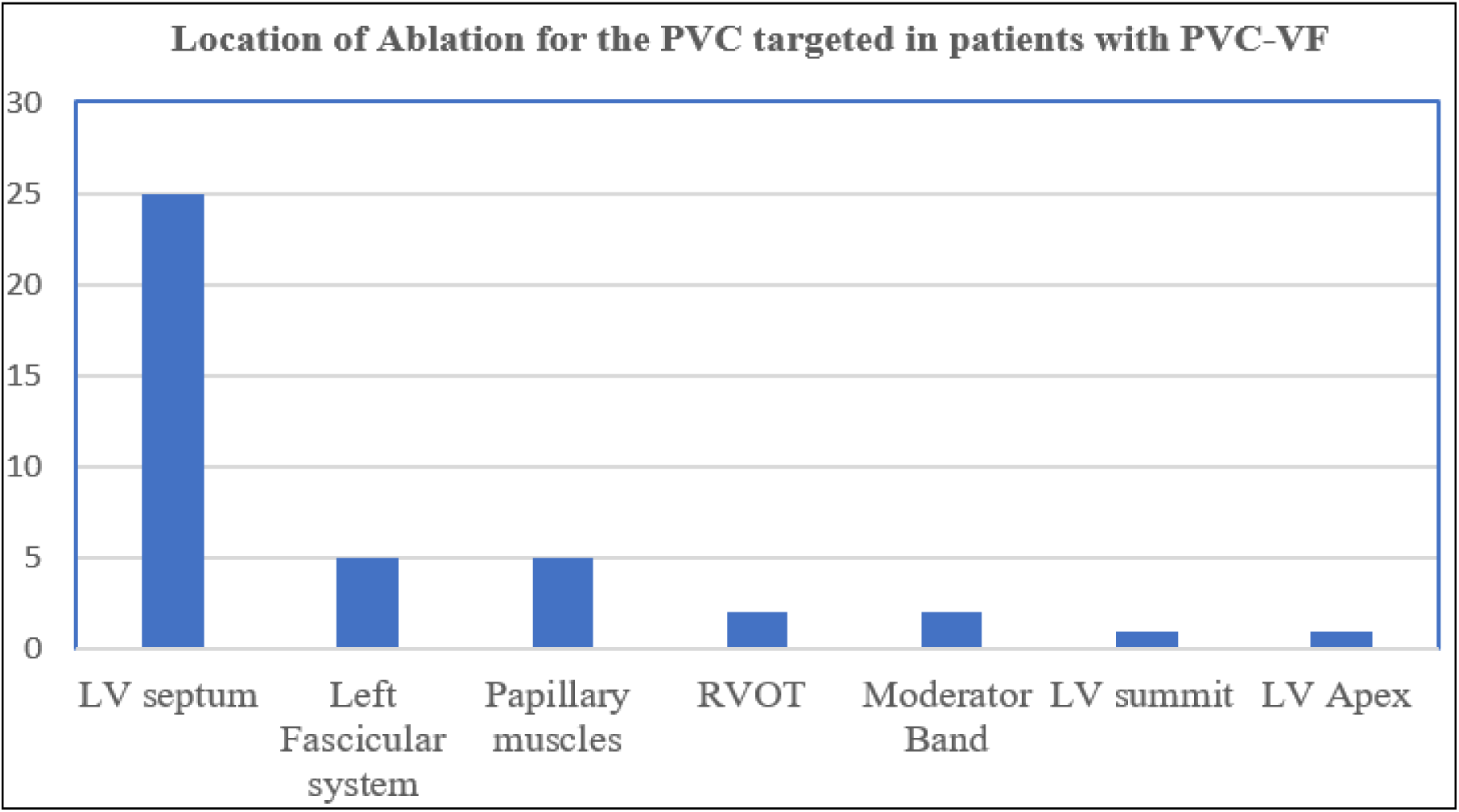
Location of ablation for PVC targeted in patients with PVC triggered VF.

**Table 2:** Ablation characteristics and Clinical Outcomes after Ablation for PVC triggered Ventricular Fibrillation.

| **EKG characteristics** | PVC-VF subgroup<br>(N = 41) | Control subgroup<br>(N = 20) | P-value |
| --- | --- | --- | --- |
| QRS duration of PVCs (ms) | 141 ± 28 | 145 ± 11.5 | 0.61 |
| Coupling interval of PVCs (ms) | 313 ± 130 | 455 ± 48.1 | 0.01 |
| Left Bundloid morphology | 5 (12%) | 10 (55%) | 0.01 |
| Right Bundloid morphology | 30 (73%) | 5 (25%) | 0.01 |
| Notching of PVC QRS complex | 29 (71%) | 5 (25%) | 0.001 |
| Early repolarization pattern | 16 (39%) | 4 (20%) | 0.001 |
| **Ablation characteristics** |  |  |  |
| Number of PVCs observed during ablation | 1.4 ± 0.9 | 1.6 ± 0.7 | 0.10 |
| VF induced during ablation | 28 (68.3%) | 0 | NA |
| Purkinje potentials targeted during ablation | 25 (60.9%) | 0 | NA |

### Deep Learning ECG Discrimination of PVC Triggered Ventricular Fibrillation

Using an auto-encoder deep learning ECG algorithm, we identified 8 key ECG ‘factors’ associated with the prediction of PVC-triggered VF as compared PVC without VF. When combining these key factors, the deep learning ECG algorithm demonstrated a reasonable discrimination between PVC triggered VF and PVC without a history of ventricular arrhythmia (AUROC 0.85 [0.56-1.0]). In explainability analysis, the two most salient ECG signatures associated with PVC-triggered VF were (1) anterior ST segment deviation and (2) left bundle branch like morphology (**Figure 5).**

**Figure 5:**
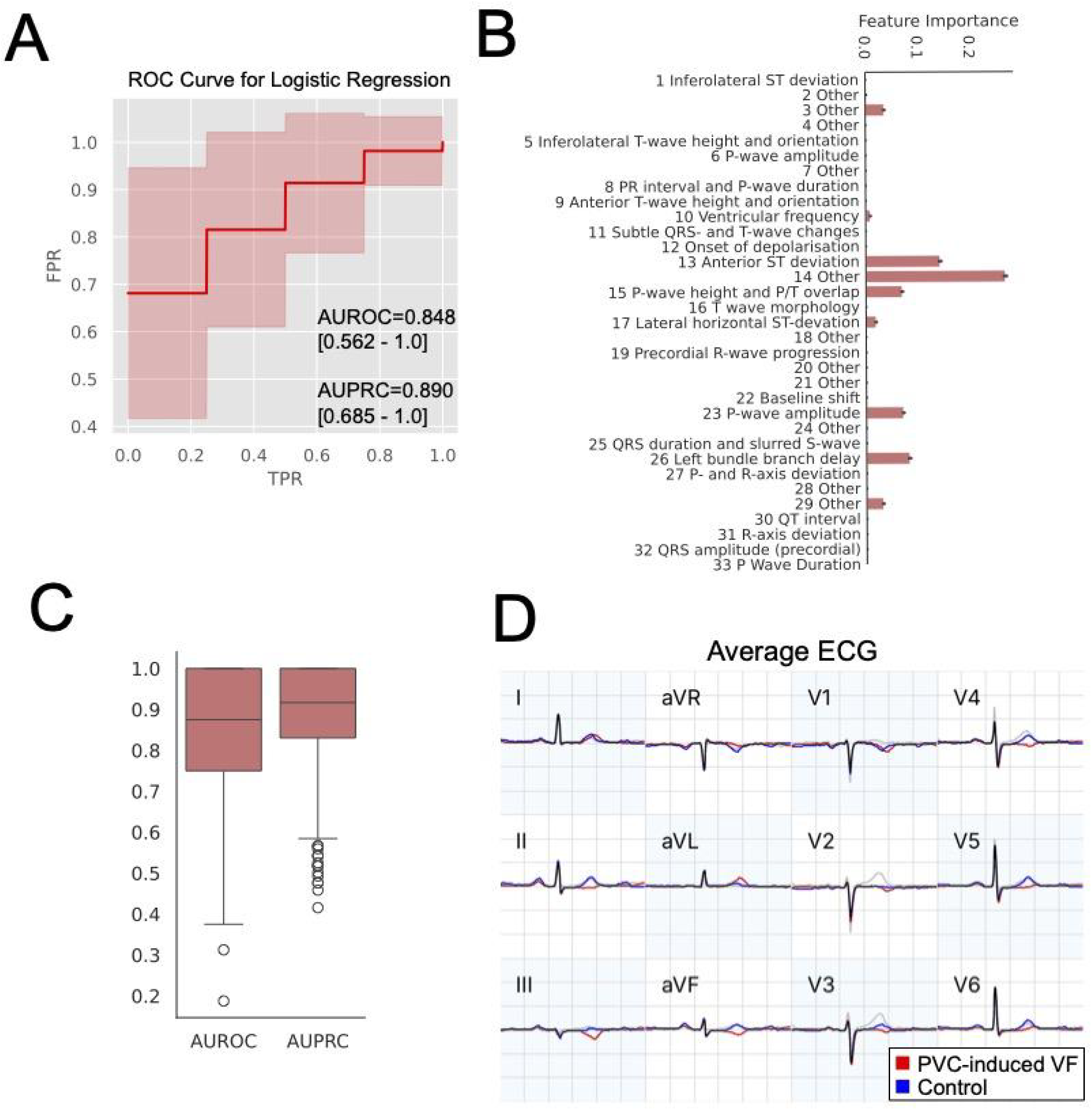
Use of FactorECG in Predicting PVC induced VF. (A) Receiver operating characteristic curve analysis for FactorECG. (B) Explainability of each individual FactorECG “factor” in prediction PVC induced VF. (C) Boxplot of area under the receiver-operating characteristic (AUROC) and area under the precision-recall curve (AUPRC). (D) Visual representation of median beat ECG differences between patients with PVC induced VF and patients undergoing ablation for PVC not inducing VF

## Discussion

In this multi-center cohort of patients with PVCs with and without VF, we show that patients with PVC-triggered VF had a higher prevalence of 12 lead EKG abnormalities including early repolarization pattern and QRS notching when compared to patients with PVCs and no history of ventricular arrhythmia. We also show that catheter ablation of trigger and substrate yielded freedom from VF in the majority of patients after median of 1 procedure. We demonstrate that a deep learning ECG algorithm applied to the sinus rhythm ECG effectively discriminated patients with PVC-triggered VF as compared to those with PVCs and no history of VF. Specific sinus rhythm ECG signatures that were associated with prediction of PVC-triggered VF were anterior ST segment deviation and left bundle branch block morphology.

While PVCs are commonly encountered in clinical practice, there is now well-established recognition that a subset of individuals with PVCs can be at risk for PVC-triggered VF and sudden death [22]. While there has been substantial evolution of our understanding of interventional approaches to this condition – including PVC ablation and denetworking of the Purkinje system – effective tools for risk stratification in the broader population of patients with PVCs are lacking [25]. Previous studies have suggested that the coupling interval between a PVC and the antecedent sinus beat could be a helpful arbiter of ventricular arrhythmia risk in those with PVCs. For example, in an observational study from Hamon and colleagues, a variable coupling interval was associated with higher risk than a fixed PVC coupling interval (either short or long) [26]. However, more recent multi-center data from the THESIS and CASPER registries have indicated risk in individuals with PVC regardless of coupling interval [5, 9]. Taken together, these conflicting data underscore the limitations of relying on coupling interval as a risk stratification tool in individuals with PVCs. To that end, there has been a parallel interest in understanding the role of myocardial substrate in the pathogenesis of PVC-triggered VF [29] and the relationship of this abnormal substrate to the triggering PVC [30]. For example, high-density electrogram mapping in patients with idiopathic VF demonstrated frequent co-localization of abnormal electrograms and drivers of VF [31]. Areas of identified electrical abnormality were often subtle and regionally small, underscoring the potential limitations of other screening methods such as cardiac magnetic resonance imaging as a risk stratification tool.

Given these potential limitations, we sought to evaluate if the 12 lead ECG – a low, cost and scalable tool – could be used to enhance risks identification in patients with PVCs. We evaluated both visually apparent ECG abnormalities as well as the utility of a deep learning algorithm incorporating ECG waveforms. First, we show that that visually apparent ECG abnormalities (inferior early repolarization, QRS notching) were significantly more commonly present in patients with PVC-triggered VF as compared to patients with PVC without VF. We hypothesize that these findings, mechanistically, are reflective of impaired electrical dynamics and repolarization heterogeneity that underlie a vulnerable substrate in patients with PVC-triggered VF. Previous studies have linked early repolarization with epicardial electrogram abnormalities generally [34] and at least one other group has similarly identified early repolarization and fractionated QRS morphology in patients with PVC-triggered VF [35].

Second, we demonstrate the role of a deep learning algorithm to distinguish patients with PVC-triggered VF and patients with PVC without VF. Previous studies have demonstrated the utility of deep learning PVC algorithms to identify patients at risk for PVC-induced cardiomyopathy, potentially identifying ECG features capturing impaired excitation-contraction coupling and autonomic dysfunction [12]. Others have also suggested that deep learning analysis of the ECG may be able to predict incident ventricular arrhythmias in patients with structural heart disease [32, 33]. Here, we show as a proof of principle, that a deep learning algorithm could distinguish between patients with PVC-triggered VF as compared to PVC without VF. In explainability analysis, we find that anterior ST segment deviation and left bundle branch like morphology as salient predictors of risk. We hypothesize, from a mechanistic standpoint, that these features may capture subtle depolarization abnormalities that may be salient to risk in those with PVC-triggered VF. Given the low-cost and scalable nature of the 12 lead ECG, if these findings can be replicated in a larger prospective evaluation, such a deep learning-based approach could transform the paradigm of malignant arrhythmia screening in individuals with PVCs.

### Limitations

We acknowledge several limitations of our study. First, this is a retrospective cohort study from an international multi-center study group. Whether or not these findings, and specifically ablation outcomes, are generalizable to other centers is not known. Second, while the assessment of ECG abnormalities was performed with blinded review and discrepancies resolved by third party assessment, we note that real-world evaluation of these abnormalities may be subject to ascertainment variability which could influence clinical utility. Third, the interventional approach to PVC-triggered VF ablation was not prospectively harmonized which could have introduced heterogeneity into the analysis of outcomes. That said, by its virtue as a real-world cohort where ablation strategy was deferred to individual operators, we believe this work may be more generalizable as it reflects the empiric heterogeneity of ablation approach in this condition. Finally, given the size and retrospective nature of this cohort, we would emphasize that these findings reflect key pilot data and that further prospective, large-scale assessment may be warranted.

### Conclusion

In summary, in this retrospective, international multi-center cohort study, we show that visually assessed abnormalities (inferior repolarization abnormality, QRS notching) and deep learning-based analysis from a routine 12 lead ECG was able to effectively distinguish patients with PVC-triggered VF from those with PVC without VF.

## Data Availability

Data will be made available based on reasonable request

## References

1. Kennedy HL, Whitlock JA, Sprague MK, Kennedy LJ, Buckingham TA, Goldberg RJ. Long-term follow-up of asymptomatic healthy subjects with frequent and complex ventricular ectopy. N Engl J Med 1985; 312: 193–197.

2. Kim YG, Choi YY, Han KD, Min KJ, Choi HY, Shim J, et al. Premature ventricular contraction increased the risk of heart failure and ventricular tachyarrhythmias. Sci Rep 2021; 11: 12698.

3. Gundalini GS, Liang JJ, Marchlinski FE. Ventricular tachycardia ablation: Past, present, and future perspectives. JACC Clin Electrophysiol 2019; 12: 1363–1383.

4. Herendael HV, Zado ES, Haqqani H, Tschabrunn CM, Callans DJ, Frankel DS, et al. Catheter ablation of ventricular fibrillation: Importance of left ventricular outflow tract and papillary muscle triggers. Heart Rhythm 2014; 11: 566–573.

5. Steinberg C, Davies B, Mellor G, Tadros R, Laksman ZW, Roberts JD, et al. Short-coupled ventricular fibrillation represents a distinct phenotype among latent causes of unexplained cardiac arrest: a report from the CASPER registry. Eur Heart J 2021; 42: 2827–2838.

6. Hu D, Viskin S, Oliva A, Carrier T, Cordeiro JM, Martinez-Barajas H, et al. Novel mutation in the SCN5A gene associated with arrhythmic storm development during acute myocardial infarction. Heart Rhythm 2007; 4: 1072–80.

7. Haissaguerre M, Derval N, Sacher F, Jesel L, Deisenhofer I, de Roy L, et al. Sudden cardiac arrest associated with early repolarization. N Engl J Med 2008; 358: 2016–23.

8. Komatsu Y, Nogami A, Hocini M, Morita H, Sato N, Marijon E, et al. Triggers of ventricular fibrillation in patients with inferolateral J-wave syndrome. J Am Coll Cardiol EP 2024; 10: 1–12.

9. Belhassen B, Conte B, Steinberg C, Whitaker J, Khan HR, Laredo M, et al. Mode and characteristics of arrhythmia initiation in idiopathic ventricular fibrillation: A THESIS substudy. JACC Clin Electrophysiol 2024; 10: 1794–1809.

10. Cheng YJ, Li ZY, Yao FJ, Xu XJ, Ji CC, Chen MM, et al. Early repolarization is associated with a significantly increased risk of ventricular arrhythmias and sudden cardiac death in patients with structural heart disease. Heart Rhythm 2017; 14: 1157–1164.

11. Brenyo A, Pietrasik G, Barheshet A, Huang DT, Polonsky B, McNitt S, et al. QRS fragmentation and the risk of sudden cardiac death in MADIT II. J Cardiovasc Electrophysiol 2012; 23: 1343–1348.

12. Lampert J, Vaid A, Whang W, Koruth J, Miller MA, Langan MN, et al. A novel ECG-based deep learning algorithm to predict cardiomyopathy in patients with premature ventricular complexes. JACC Clin Electrophysiol 2023; 9: 1437–1451.

13. Tigen K, Karaahmet T, Gurel E, Cevik C, Nugent K, Pala S, et al. The utility of fragmented QRS complexes to predict significant interventricular dyssynchrony in nonischemic dilated cardiomyopathy patents with narrow QRS interval. Can J Cardiol 2009; 25: 517–522.

14. Ahn MS, Kim JB, Joung B, Lee MH, Kim SS. Prognostic implications of fragmented QRS and its relationship with delayed contrast-enhanced cardiovascular magnetic resonance imaging in patients with non-ischemic dilated cardiomyopathy. Int J Cardiol 2013; 167: 1417–1422.

15. Das MK, Khan B, Jacob S, Kumar A, Mahenthiran J. Significance of a fragmented QRS complex versus a Q wave in patients with coronary artery disease. Circulation 2006; 113: 2495–2501.

16. Macfarlane PW, Antzelevitch C, Haissaguerre M, Huikuri HV, Potse M, Rosso R, et al. The early repolarization pattern: A consensus paper. J Am Coll Cardiol 2015; 66:

17. Van de Leur RR, de Brouwer R, Bleijendaal H, Verstraelen TE, Mahmoud B, Perez-Matos A, et al. ECG-only explainable deep learning algorithm predicts the risk of malignant ventricular arrhythmia in phospholamban cardiomyopathy. Heart Rhythm 2024; 21: 1102–1112.

18. Van de Leur RR, Bos MN, Taha K, Sammani A, Yeung MW, van Duijvenboden S, et al. Improving explainability of deep neural network-based electrocardiogram interpretations using variational auto-encoders. Eur Heart J Digit Health 2022; 3: 390–404.

19. Nogami A, Sugiyasu A, Kubota S, Kato K. Mapping and ablation of idiopathic ventricular fibrillation from the Purkinje system. Heart Rhythm 2005; 2: 646–649.

20. Prisco AR, Castro JR, Roukoz H, Tholakanhalli VN. Premature ventricular complexes: Benign versus Malignant-How to approach? Indian Pacing Electrophysiol J 2023; 23: 189–195.

21. Niwano S, Wakisaka Y, Niwano H, Fukaya H, Kurokawa S, Kiryu M, et al. Prognostic significance of frequent premature ventricular contractions originating from the ventricular outflow tract in patients with normal left ventricular function. Heart 2009; 95: 1230–1237.

22. Haissaguerre M, Shoda M, Jais P, Nogami A, Shah DC, Kautzner J, et al. Mapping and ablation of idiopathic ventricular fibrillation. Circulation 2002; 106: 962–967.

23. Santoro F, Di Biase L, Hrantizky P, Sanchez JE, Santangeli P, Perini AP, et al. Ventricular fibrillation triggered by PVCs from papillary muscles: clinical features and ablation. J Cardiovasc Electrophysiol 2014; 25: 1158–64.

24. Sadek MM, Benhayon D, Sureddi R, Chik W, Santangeli P, Supple GE, et al. Idiopathic ventricular arrhythmias originating from the moderator band: electrocardiographic characteristics and treatment by catheter ablation. Heart Rhythm 2015; 12: 67–75.

25. Salazar P, Beaser AP, Upadhyay GA, Aziz Z, Besser S, Shatz DY, et al. Empiric ablation of polymorphic ventricular tachycardia/fibrillation in the absence of a mappable trigger: Prospective feasibility and efficacy of pacemap matching to defibrillator electrograms. Heart Rhythm 2022; 19: 527–33.

26. Hamon D, Rajendran PS, Chui RW, Ajijola OA, Irie T, Talebi R, et al. Premature ventricular contraction coupling interval variability destabilizes cardiac neuronal and electrophysiological control: Insights from simultaneous cardioneuronal mapping. Circ Arrhythm Electrophysiol 2017; 10: e004937.

27. Cluitmans MJM, Bear LR, Nguyen UC, van Rees B, Stoks J, Ter Bekke RMA, et al. Noninvasive detection of spatiotemporal activation-repolarization interactions that prime idiopathic ventricular fibrillation. Sci Transl Med 2021; 13: eabi9317.

28. Walton RD, Martinez ME, Bishop MJ, Hocini M, Haissaguerre M, Plank G. Influence of the Purkinje-muscle junction on transmural repolarization hetereogeneity. Cardiovasc Res 2014; 103: 629–640.

29. Haissaguerre M, Duchateau J, Dubois R, Hocini M, Cheniti G, Sacher F, et al. Idiopathic ventricular fibrillation and microstructural abnormalities. JACC: Clinical Electrophysiol 2020; 6: 591–608.

30. Baskovski E, Altin T, Akyurek O, Tan TS, EkremCunetoglu M, Akbulut IM, et al. Fascicular/Purkinje tissue colocalized with scar in cardiomyopathy patients undergoing ventricular fibrillation ablation. Pacing Clin Electrophysiol 2025; 48: 672–681.

31. Haissaguerre M, Hocini M, Cheniti G, Duchateau J, Sacher F, Puyo S, et al. Localized structural alterations underlying a subset of unexplained sudden cardiac death. Circ Arrhythm Electrophysiol 2018; 11: e006120.

32. Oberdier MT, Neri L, Orro A, Carrick RT, Nobile MS, Jaipalli S, et al. Sudden cardiac arrest prediction via deep learning electrogram analysis. Eur Heart J Digit Health 2025; 6: 170–179.

33. Taye GT, Shim EB, Hwang J-H, Lim MK. Machine learning approach to predict ventricular fibrillation based on QRS complex shape. Front Physiol 2019; 10: 1193.

34. Zhang J, Hocini M, Strom M, Cuculich PS, Cooper DH, Sacher F, et al. The electrophysiological substrate of early repolarization syndrome: Noninvasive mapping in Patients. JACC Clic Electrophysiol 2017; 3: 894–904.

35. Arceluz MR, Thind M, Garcia FC, Guandilini GS, Santangeli P, Hyman M, Deo R, et al. Sinus rhythm electrocardiographic abnormalities, sites of origin, and ablation outcomes of ventricular premature depolarization initiating ventricular fibrillation. Heart Rhythm 2023; 20: 844–852.

36. Tikkanen JT, Annttonen O, Junttila MJ, Aro AL, Kerola T, Rissanen HA, et al. Long-term outcome associated with early repolarization on electrocardiography. N Engl J Med 2009; 361: 2529–2537.

37. Sinner MF, Reinhard W, Muller M, Beckmann BM, Martens E, Perz S, et al. Association of early repolarization pattern on ECG with risk of cardiac and all-cause mortality: a population-based prospective cohort study (MONICA/KORA). PloS Med 2010; 7: e1000314.

38. Salazar P, Beaser AP, Upadhyay GA, Aziz Z, Besser S, Shatz DY, et al. Empiric ablation of polymorphic ventricular tachycardia/fibrillation in the absence of a mappable trigger: Prospective feasibility and efficacy of pacemap matching to defibrillator electrograms. Heart Rhythm 2022; 19: 527–33.

39. Lowery CM, Tzou WS, Aleong RG, Nguyen DT, Varosy PD, Katz DF, et al. Use of stored implanted cardiac defibrillator electrograms in catheter ablation of ventricular fibrillation. Pacing Clin Electrophysiol 2013; 36: 76–85.

40. Nakamura T, Schaeffer B, Tanigawa S, Muthalay RG, John RM, Michaud GF, et al. Catheter ablation of polymorphic ventricular tachycardia/fibrillation in patients with and without structural heart disease. Heart Rhythm 2019; 16: 1021–1027.

